# Ethnic Disparities in Acute Stroke Presentation and Reperfusion Therapy in a Dutch Comprehensive Stroke Center

**DOI:** 10.64898/2026.04.23.26351631

**Authors:** Y.X. Lee, P.V. Hurkmans, H.J. Arwert, T.P.M. Vliet Vlieland, I.R. van den Wijngaard, D.E.C.M. Hofs, K. Jellema

## Abstract

**Objective:** To assess ethnic disparities in time to hospital presentation, use of acute reperfusion therapies, and in-hospital treatment times among patients presenting with stroke in a Dutch emergency department.

**Methods:** In this single-centre observational cohort study, we included patients with a first-ever ischemic stroke between September 2020 and September 2021. Patients were categorized by ethnicity (with or without migration background). Demographic and stroke characteristics were compared between groups. Outcomes included: rates of presentation outside therapeutic time window, acute reperfusion therapy (intravenous thrombolysis (IVT) and endovascular thrombectomy (EVT)), and, when applicable, door-to-treatment time (DTTT), with a door-to-needle time (DTNT) and door-to-groin time (DTGT) for IVT and EVT respectively. Univariable and multivariable linear and logistic regression analyses were performed, adjusted for age, sex, and NIHSS at presentation, where appropriate.

**Results:** A total of 232 patients were included, of whom 62 (26.7%) had a migration background. These patients were younger (66.6 vs 71.2 years) and more frequently had diabetes (27.4% vs 15.9%). Sex distribution was similar (59.7% vs 60.6% male). Stroke etiology differed between groups with less cardio-embolism (4.8% vs 15.3%) and more small vessel disease (69.4% vs 48.2%) among patients with a migration background. These latter patients presented more often outside the therapeutic time window (53.2% vs 37.1%; OR 1.90; 95% CI 1.05-3.45). EVT was less frequently performed in patients with a migration background compared to those without (8.1% vs 22.4%; OR 0.28; 95% CI 0.10-0.75). There were no significant differences in treatment times (DTTT 38min vs 30min, DTNT 35min vs 26min, DTGT 64min vs 54min).

**Conclusion:** Patients with a migration background were more likely to present outside the therapeutic time window and had a lower rate of EVT. In order to improve access for these patients, more insight into prehospital and within hospital barriers and facilitators for appropriate management are needed.

## INTRODUCTION

In the Netherlands, approximately 42.000 persons experienced a new stroke in 2024. About 80% of these patients have ischemic stroke.(1,2) In many stroke survivors, stroke causes long-term disability within several domains of health, with a significant negative impact on the long-term health-related quality of life.(3–6) These long-term disability and reduced quality of life following stroke contribute to significant societal impact and costs.(7) With respect to ischemic stroke, timely recognition of symptoms is important to start acute reperfusion therapy as soon as possible, as previous studies have shown that earlier treatment leads to a better functional outcome and quality of life.(8,9) Reperfusion therapy for ischemic stroke consists of intravenous thrombolysis (IVT) and endovascular thrombectomy (EVT).

Over the past years, literature has shown that there are disparities in acute stroke care. Factors associated with the timely and adequate institution of acute treatment were found to be diverse, including geographical location, socioeconomic status, sex, and also ethnicity. (10–12) Ethnicity is a social-political construction and is defined as a group of people that has a common national or cultural tradition.(13) A systematic review investigating the role of ethnicity in stroke care included 30 studies, all conducted in the US.(11) The authors concluded that white patients were more likely to arrive at the emergency department within three hours from onset of stroke symptoms compared to Black and Hispanic patients. Additionally, Black, Hispanic, and Asian patients were less likely to receive acute reperfusion therapy compared to white patients.(11) These findings are in line with those of a recent meta-analysis examining multiple determinants of acute stroke care, which included 38 studies, primarily from the US and New Zealand.(10) Within these 38 studies, 14 studies assessed ethnicity in relation to IVT rate and 11 studies in relation to EVT rate.(10) The authors found that Black patients had lower rates of acute reperfusion therapy compared to white patients.

These reviews did not include European studies. Since the publication of these reviews, three additional observational studies have been published (14–16), including a European study conducted in the UK (16), again showing lower acute reperfusion therapy rates and delayed hospital presentation among Black patients. Whereas the occurrence of ethnic disparities in stroke care in the Netherlands is not unlikely, studies regarding the relation between ethnicity and stroke presentation, stroke recognition and acute treatment in the Netherlands are scarce. There is one study in the Netherlands from 2011 (17), which was excluded from aforementioned meta-analysis (10) due to its publication date. This study found that non-white patients were less likely to receive IVT than white patients, partly due to delayed hospital presentation.(17) Observed differences by ethnicity may in part reflect inequities in access to care, differences in healthcare utilization, and potential implicit bias in clinical decision-making. Over the past decade, acute stroke treatment has evolved substantially. Since the publication of the MR CLEAN trial in 2015 (18), which showed that EVT added to standard care significantly improved functional outcomes in patients with stroke caused by large vessel occlusion, EVT has become an established treatment option, and the eligibility criteria for IVT have expanded. (18,19) It remains unclear whether previously observed ethnic disparities persist with current management.

To reduce inequity in stroke care management, it is important to detect these disparities to create interventions targeting specific high-risk groups. Therefore, the current study aims to investigate the association between ethnicity and timing of hospital presentation, use of acute reperfusion therapy, and in-hospital treatment times among patients with stroke presenting on the emergency department in the Netherlands.

## METHODS

### Study design

This study used data from a prospective single-center cohort study that included patients from September 2020 to September 2021. The design and data collection of this study have been described previously in detail.(20) Briefly, patients were included at the comprehensive stroke center Haaglanden Medical Center (HMC) in The Hague, a multicultural city in the Netherlands. Data were obtained from patients’ medical records and a digital questionnaire that was administered at admission and at 3– and 12-months follow-up. A written exemption from ethical approval was acquired from the Medical Ethics Review Committee South West Netherlands (METC nr: Z19.039). The study was conducted according to the Declaration of Helsinki.(21) For the present analysis, only a subset of data (no follow-up data) from the larger study were used. The reporting is done according to The Strengthening the Reporting of Observational Studies in Epidemiology (STROBE) guidelines.(22)

### Study population

We included patients aged ≥18 years with a first-ever ischemic stroke who were seen on the emergency department of the hospital and who were hospitalized subsequently. Exclusion criteria were: hemorrhagic stroke, subarachnoid hemorrhage, traumatic intracerebral hemorrhage or venous sinus thrombosis. Patients who were unable to complete questionnaires in Dutch (e.g. due to an insufficient understanding of Dutch, severe cognitive problems or severe aphasia) were also excluded. One researcher (Y.X.L.) screened patients from the larger study database to identify those eligible for the present study.

### Measurements

Assessments included extraction of data from patients’ medical records as well as administration of a digital questionnaire at admission. One researcher (Y.X.L.) extracted data on stroke characteristics, data of the presentation at the emergency department, and treatment details from the medical records. A research nurse collected the questionnaires for sociodemographic characteristics.

#### Sociodemographic and stroke characteristics

##### Sociodemographic characteristics

The following sociodemographic characteristics were gathered: age (in years); sex (male/female); educational level as a proxy variable data for socioeconomic status (low: up to and including lower technical and vocational training; medium: up to and including secondary technical and vocational training; high: up to and including higher technical and vocational training and university); working before stroke (yes/no); alcohol utilization before event (yes/no); smoking before event (yes/no); oral anticoagulation use (yes/no); body mass index (BMI); having comorbidities (it was possible for patients to mark more than one answer with a choice of: transient ischemic attack /myocardial infarction/arrhythmia/diabetes mellitus/hypertension/hypercholesterolemia/none).

##### Ethnicity

In this study, we categorized patients as a group of patients with a migration background and a group of patients without a migration background. Regarding migration background, we used the definition provided by the Statistics Netherlands (23): Patients were considered to have a migration background if they were born outside the Netherlands or had at least one parent born outside the Netherlands.(23) The country of birth of patient and parent(s) were obtained via the baseline questionnaire.

##### Stroke characteristics

The following stroke characteristics were extracted from the patients’ medical records: National Institutes of Health Stroke Scale (NIHSS) score at admission; etiology of stroke according Trial of Org 10172 in Acute Stroke Treatment (TOAST) (large-artery atherosclerosis/cardio embolism/small-vessel disease/other determined cause/undetermined cause); stroke side (left/right); affected circulation (anterior/posterior); in case of a large vessel occlusion, the location of the occlusion (intracranial segment of the internal carotid artery/terminal segment of the internal carotid artery/middle cerebral artery first segment/middle cerebral artery second segment proximal or distal/basilar artery/other); recanalization in case of EVT with a Thrombolysis in Cerebral Infarction score (TICI) ≥2B; complications of the acute reperfusion treatment (yes/no).

#### Outcomes

All outcome data were collected from patients’ medical records. The primary outcomes were timing of presentation to the emergency department and use of any form of acute reperfusion therapy. For timing of presentation, we calculated the proportion of patients presenting outside the therapeutic time window. Use of acute reperfusion therapy was further divided into IVT and EVT. Additional outcomes included the door-to-treatment time (DTTT) among patients who received acute reperfusion therapy. This outcome was further specified as door-to-needle time (DTNT) for patients who received IVT and door-to-groin time (DTGT) for those who underwent EVT. In patients who received both therapies, DTTT was calculated based on the first treatment administrated. In addition, the prevalence of contraindications for acute reperfusion therapy were documented, as determined by the neurologist at the time of assessment and documented in the medical record. The contraindications were categorized by: use of oral anticoagulation, blood pressure ≥ 185/110 mmHg, non-disabling neurological deficits and other (i.e. recent major operation or relevant extracranial bleeding).

### Data analysis

All statistical analyses were conducted using SPSS Statistics, version 29 (Armonk, NY: IBM Corp). Baseline characteristics were analyzed using descriptive statistics, with continuous data presented as mean with standard deviation (SD) or median with interquartile range, depending on their distribution, whereas for categorical data numbers and percentages were calculated. To compare the baseline characteristics of patients with and without a migration background, unpaired-t tests, Mann-Whitney U tests or Chi-Square tests were used, where appropriate. Differences in certain stroke etiology might contribute in eligibility for EVT, as large-artery atherosclerosis and cardio-embolism are main causes for large vessel occlusion, which is treated with EVT.(24) Therefore, if significant differences between groups were found with the Chi-square test, a post-hoc analysis for the variable stroke etiology was performed to identify which etiology contributed most to the observed differences. This was assessed using adjusted standardized residuals of the chi-square test, with residuals with an absolute value greater than 1.96 being considered to indicate a statistically significant deviation from expected frequencies.

To examine the association between migration background and the various outcomes, we performed univariable and multivariable logistic regression analyses with calculation of the odds ratios (OR) with the 95% Confidence Interval (CI) for the binary outcomes (utilization of any reperfusion treatment subdivided by IVT and EVT, presentation outside therapeutic time window and other contraindications of acute reperfusion therapy). All multivariable analyses were adjusted for age and sex. To avoid model overfitting in the multivariable logistic regression analyses due to a small number of events and group imbalance, a limited set of additional predetermined covariates (NIHSS at presentation, comorbidity diabetes mellitus, comorbidity hypertension, use of oral anticoagulation, level of education) was considered for statistical adjustment. Eventually, those variables were only included if their separate inclusion resulted in a ≥10% change in the OR compared to the unadjusted model, as this would suggest of possible confounding.(25) If the number of events in a group was <5, instead of a logistic regression analysis, we performed a Fisher’s exact test, as OR is then unreliable and adjustment for confounding variables is not possibility.(26–28)

For the in-hospital treatment times (DTTT, DTNT and DTGT), we performed both a univariable and multivariable linear regression analysis. The multivariable linear models were adjusted for sex, age, and NIHSS at presentation. This set of covariates was included as, compared to the logistic regression models, linear regression models generally are less constrained by sparse outcome data and are primarily limited by the total sample size rather than the number of events.(29,30) Unadjusted and adjusted beta coefficients (B) with the 95% CI were reported. In cases of non-normally distributed residuals of the linear regression analysis, data were log-transformed using the natural logarithm prior to linear regression analysis. Analyses were performed using complete cases, observations with missing values on variables included in the aforementioned models were excluded. The proportion of missing data per variable is presented in each table. Given the low expected proportion of missing data, imputation was not planned. A two-sided p-value of ≤0.05 was considered statistically significant.

## RESULTS

### Patient characteristics

Between September 2020 to September 2021, a total of 475 patients were considered eligible for participation for the larger study including consecutive patients with an ischemic or hemorrhagic stroke. Of those, 263 patients (55.4%) provided informed consent. Regarding the selection of patients for the present analysis, one patient was excluded due to lack of information about ethnicity. Thirty patients were excluded due to a hemorrhagic stroke as diagnosis. Finally, a total of 232 patients with an acute ischemic stroke were included, of whom 62 (26.7%) had a migration background and 170 (73.3%) did not.

Baseline characteristics categorized and compared according to migration background are presented in Table 1. Patients with a migration background were significantly younger (66.6 vs 71.2 years, p=0.02). The sex distribution in patients with a migration background was similar compared to patients without a migration background. Patients with a migration background more often had diabetes mellitus compared to patients without a migration background. Patients with a migration background also more often were smokers, although this difference was not statistically significant. The distribution of stroke etiology differed significantly according to migration background (p=0.02). Therefore, a post-hoc analysis of the adjusted standardized residuals of the Chi-square test was performed, which indicated that small-vessel disease occurred more frequently than expected among patients with a migration background (69.4% vs 47.6%; adjusted standardized residual = 2.9). In contrast, cardio-embolic strokes occurred less frequently than expected in this group (4.8% vs 15.3%; adjusted standardized residual = −2.1). The proportion of large artery atherosclerotic strokes was also lower than expected (6.5% vs 15.9%; adjusted standardized residual = –1.9), although this did not reach statistical significance. There were no statistically significant differences in NIHSS scores, stroke location, recanalization rate after EVT or complication rate after treatment among both groups. The proportion of missing data per baseline variable was low (<5%) and is presented in the footnote of Table 1.

**Table 1.**
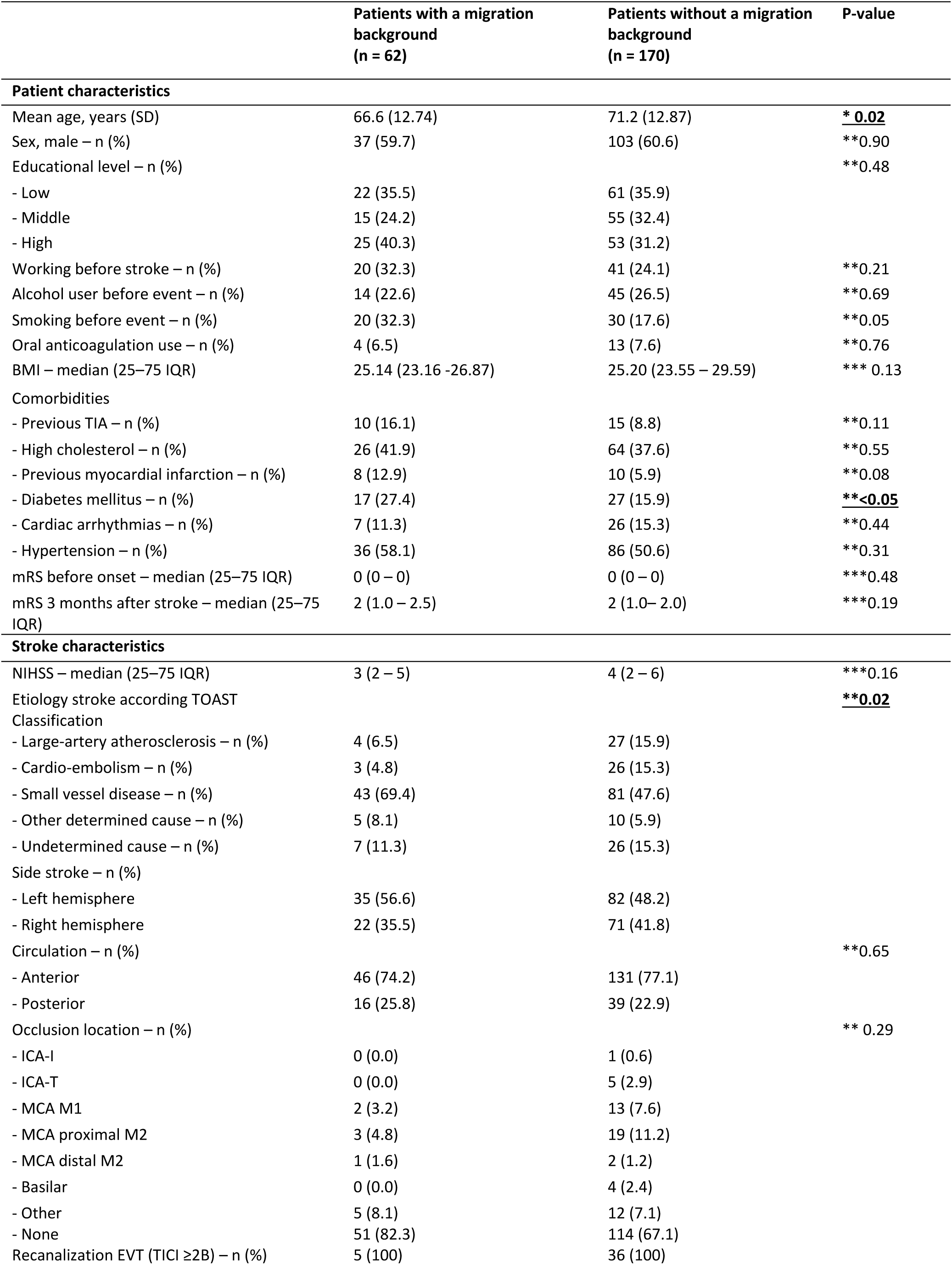
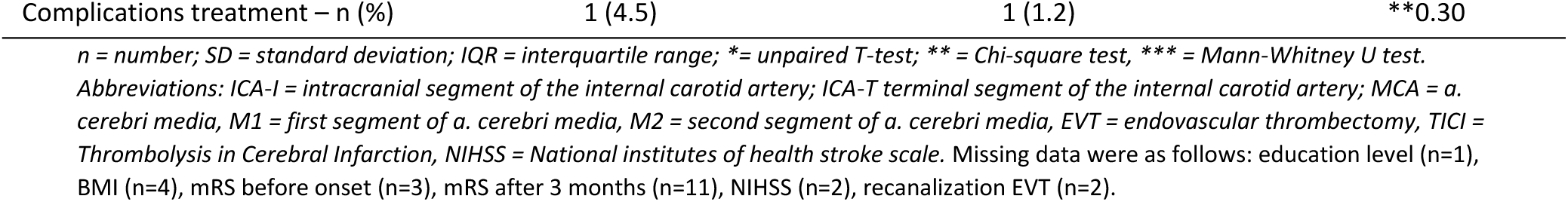
Baseline table; differences between patients with a migration background and patients without a migration background.

### Eligibility for reperfusion therapy

Results of the rates of presentation outside therapeutic time window and contraindications in patients with and without a migration background are presented in Table 2. No missing data were present for these variables. Patients with a migration background more frequently presented outside the therapeutic time window compared with patients without a migration background (53.2% vs 37.1%, adjusted OR 1.90, 95%-CI 1.05-3.45, p=0.03). The presence of non-disabling neurological deficit as a contraindication for treatment did not significantly differ between groups. For both outcomes, presentation outside the therapeutic time window and non-disabling neurological deficits as contraindication, statistical adjustment was performed only for age and sex, as adding aforementioned predetermined covariates did not result in ≥10% change in the OR compared with the unadjusted model. No statistically significant differences were observed for the impact of other causes of ineligibility for acute reperfusion therapy.

**Table 2.** Results rates contra indications and treatment acute reperfusion therapy with univariate and multivariate logistic regression analysis.

|  | PMB+<br>(n = 62) | PMB-<br>(n = 170) | Unadjusted OR<br>(95% CI) | P-value | Adjusted OR<br>(95% CI) | P-value |
| --- | --- | --- | --- | --- | --- | --- |
| **Not eligible for acute reperfusion therapy** |  |  |  |  |  |  |
| Presentation outside therapeutic time window – n (%) | 33 (53.2) | 63 (37.1) | 1.93 (1.07-3.48) | **0.03** | 1.90 (1.05-3.45)* | **0.03** |
| Contraindication – n (%) |  |  |  |  |  |  |
| - Oral anticoagulation | 0 (0) | 3 (1.8) | n/a | **0.57 | n/a | n/a |
| - Blood pressure ≥ 185/110 mmHg | 1 (1.6) | 0 (0) | n/a | **0.27 | n/a | n/a |
| - Non-disabling neurological deficits | 5 (8.1) | 15 (8.8) | 0.97 (0.33-2.80) | 0.95 | 0.90 (0.31-2.65)* | 0.85 |
| - Other contraindication<sup>&</sup> | 1 (1.6) | 5 (2.9) | n/a | **1.00 | n/a | n/a |
| **Acute reperfusion therapy** |  |  |  |  |  |  |
| Any reperfusion therapy – n (%) | 22 (35.5) | 84 (49.4) | 0.56 (0.31-1.03) | 0.06 | 0.62 (0.32-1.20)<sup>#</sup> | 0.15 |
| Intravenous thrombolysis – n (%) | 20 (32.3) | 65 (38.2) | 0.77 (0.42-1.42) | 0.40 | 0.82 (0.43-1.55)<sup>#</sup> | 0.54 |
| Endovascular thrombectomy – n (%) | 5 (8.1) | 38 (22.4) | 0.31 (0.11-0.81) | **0.02** | 0.28 (0.10-0.75)* | **0.01** |

*n* = number, NIHSS = National Institutes of Health Stroke Scale, PMB+ = patients with a migration background, PMB- = patients without a migration background, IVT = intravenous thrombolysis, EVT = endovascular thrombectomy, OR = odds ratio where patients without a migration background was the reference group, CI = confidence interval, <sup>&</sup> = e.g. recent major operation or active extracranial bleeding, \* = adjusted for sex and age, <sup>#</sup> = adjusted for sex, age and NIHSS at presentation, \*\* = Fisher's exact test, n/a = not applicable.

### Provision of reperfusion therapy

Table 2 also shows the results of the provision of reperfusion therapies. A total of 106 patients received a form of acute reperfusion therapy, with no missing data. Patients with a migration background less often received any form of acute reperfusion therapy (35.5% vs 49.4%), although this was not statistically significant (unadjusted OR: 0.56, 95% CI: 0.31-1.03, p=0.06; adjusted OR: 0.62, 95% CI: 0.32-1.20, p=0.15). Patients with a migration background had a significantly lower rate of undergoing EVT compared to patients without a migration background (8.1% vs 22.4%, unadjusted OR: 0.31, 95% CI: 0.11–0.81, p=0.02; adjusted OR: 0.28, 95% CI: 0.10–0.75, p=0.01). No difference was observed for treatment with IVT (32.3% vs 38.2%, unadjusted OR: 0.77, 95% CI: 0.42–1.42, p=0.40; adjusted OR: 0.82, 95% CI: 0.43–1.55, p=0.54). For the multivariable logistic regression analyses of any reperfusion therapy and IVT, adjustment was performed for age and sex, and additionally for NIHSS at presentation, as inclusion of NIHSS resulted in a ≥10% change in the OR compared with the unadjusted model. Other predefined covariates (diabetes mellitus, hypertension, use of oral anticoagulation, and level of education) did not meet this criterion and were therefore not included.

### In-hospital treatment times

In-hospital treatment times for both groups are represented in Table 3. There were no missing data for these variables. Because the residuals of the linear regression analysis of the in-hospital treatment times were not normally distributed, treatment times were log-transformed using the natural logarithm prior to analysis. Median DTTT, including DTNT and DTGT were longer for patients with a migration background compared with patients without a migration background, although these differences were not statistically significant.

**Table 3.** Results treatment times with univariate and multivariate linear regression analysis.

|  | PMB+<br>(n = 62) | PMB-<br>(n = 170) | B (95% CI) | P-value | Adjusted B*<br>(95% CI) | P-value |
| --- | --- | --- | --- | --- | --- | --- |
| DTTT (min) median (25-75 IQR) | 38.00 (21.00-60.00) | 30.00 (20.00-54.00) | 1.19 (0.87-1.63) | 0.26 | 1.17 (0.86-1.61) | 0.31 |
| DTNT (min) median (25-75 IQR) | 35.50 (20.50-54.50) | 26.00 (19.00-34.50) | 1.23 (0.92-1.64) | 0.16 | 1.19 (0.90-1.57) | 0.22 |
| DTGT (min) median (25-75 IQR) | 64.00 (54.00-84.00) | 54.00 (47.00-79.50) | 1.35 (0.76-2.40) | 0.30 | 1.39 (0.77-2.49) | 0.26 |

*PMB+ = patients with a migration background, PMB- = patients without a migration background, DTTT= door-to-treatment time, DTNT=* *door-to-needle time, DTGT= door-to-groin time, IQR = interquartile range, B = beta coefficient, CI = confidence interval, \* = adjusted for sex,* *age and NIHSS at presentation.*

## DISCUSSION

This observational study, conducted in a Dutch comprehensive stroke center, found that patients with a migration background had an almost twofold increased likelihood of presenting outside the therapeutic time window for acute reperfusion therapy for ischemic stroke in the emergency department. Moreover, EVT was less frequently performed in patients with a migration background, whereas in-hospital treatment times did not differ significantly.

The more frequent presentation outside the therapeutic time window is in line with the conclusion of a systematic review of studies conducted in the US, which reported that Black and Hispanic patients were more likely to arrive later on the emergency department after symptom onset compared to white patients.(11) The same holds for previous European studies, with a recent UK study and a 2011 Dutch study both demonstrating a higher rate of delayed presentation on the emergency department in non-white patients.(16,17) Regarding potential explanations for these observed differences, evidence from the US suggests that disparities also exist in the prehospital phase, contributing to delayed presentation and, consequently, reduced eligibility for acute reperfusion therapy.(31–33) Reported barriers include lack of awareness of the seriousness of the symptoms, previous negative experiences in accessing health care leading to avoidance, distrust in the health care system and concerns about the costs.(33) While these findings may not be directly applicable to the Dutch healthcare context, similar mechanisms may partly explain why patients with a migration background more frequently present outside the therapeutic time window.

Our finding that patients with a migration background receive EVT less frequently aligns with international evidence, primarily from the US and New Zealand, which has reported similar findings.(10,11). To date, however, no European studies have addressed this topic. Differences in EVT rates could partly be explained by the differences in presentation timing. However, differences in stroke etiology between groups may also contribute to the lower EVT rate. In our cohort, large-artery atherosclerosis and cardio-embolism were more frequently observed as underlying stroke mechanism among patients without a migration background. These etiologies are the main causes of large vessel occlusion, which is typically treated with EVT(24,34,35), whereas large vessel occlusion is uncommon in small-vessel disease as underlying etiology.(35) These findings are consistent with previous Dutch research reporting a lower prevalence of large-artery atherosclerosis among Black and Asian patients compared with white Dutch patients, and a higher prevalence of small-vessel disease among non-white Dutch patients.(36) Furthermore, a study from South-London demonstrated a higher likelihood of cardio-embolism among white patients compared to non-white patients.(37) In contrast to EVT, eligibility for IVT is not determined by the presence of a large vessel occlusion. In contrast to our results, previous European studies reported lower IVT rates among non-white patients.(16,17) One possible explanation for the absence of these differences in our study relates to the composition of our study population. Patients needed sufficient functional and cognitive capacity to complete questionnaires, resulting in the exclusion of patients with significant language barriers. Language barriers may contribute to a vulnerable situation to access of acute reperfusion therapy and health care.(38,39) Dutch statistics indicate that approximately 10% of migrants in the Netherlands speak little or no Dutch.(40)

We observed no differences in in-hospital treatment times between groups. A review of US-based studies demonstrated that Black and Hispanic patients experienced longer in-hospital treatment times compared to white patients.(11) To the best of our knowledge, there are not European studies focusing on this topic. One possible explanation for these conflicting result, is that several potential factors that may contribute to such disparities —such as healthcare accessibility, insurance coverage, and geographical distance to the hospital(11,42) —are less likely to play a substantial role within the Dutch healthcare context. In the Netherlands, all residents are legally required to obtain basic health insurance, making it less likely that individuals refrain from seeking medical care due to lack of health insurance coverage.(43) Furthermore, the Netherlands is geographically compact compared to the US. Dutch Statistics showed only limited regional variation in urgent EMS responses attributable to geographical differences.(44) Also, previous studies have demonstrated that lower socioeconomic status is associated with reduced quality stroke care.(45) In our cohort, we found no significant differences between educational level between groups, a proxy for socioeconomic status.

A strength of our study is that, to our knowledge, it is the first Dutch study focusing on ethnic disparities and acute reperfusion therapy, including EVT and in-hospital treatment times. Several limitations should also be acknowledged. Firstly, patients with significant language barriers and those with severe neurological deficits were excluded. This may have led to selection bias and underrepresentation of patients with large vessel stroke, especially in the group of patients with a migration background. Secondly, we had group imbalance and relatively low rate for the outcomes, particularly for EVT. Although adjustment for all predetermined possible confounders would have been methodologically preferable, we decided to limit the number of variables in the multivariable regression analysis model to avoid model overfitting. Therefore, it is plausible that not all relevant confounders were fully accounted for in the analysis.

In conclusion, we found ethnic disparities in the timing of hospital presentation and use of acute reperfusion therapy among patients with stroke presenting on the emergency department in the Netherlands. Future studies should adopt a multicenter design and apply broader inclusion criteria (such as the absence of restriction related to language barrier) to confirm these findings. In addition, more insight into prehospital and within hospital barriers and facilitators for appropriate management are needed in order to improve access for these patients.

**Statistical analysis completed by:** Y.X. Lee, P.V. Hurkmans

There are no “supplementary data” to this manuscript.

## Author contributions

Drafting/revising the manuscript for content, including medical writing for content: Y.X. Lee, P.V. Hurkmans, I.R. van den Wijngaard, T.P.M. Vliet Vlieland, H. Arwert, K. Jellema

Study concept or design: K. Jellema, I.R. van den Wijngaard, H. Arwert

Analysis or interpretation of data: Y.X. Lee, P.V. Hurkmans, D.E.C.M. Hofs, K. Jellema, H. Arwert

## Disclosures

Y.X. Lee reports no disclosures

P.V. Hurkmans reports no disclosures

H. Arwert reports no closures

T.P.M. Vliet Vlieland reports no disclosures

I.R. van den Wijngaard reports no disclosures

D.E.C.M. Hofs reports no disclosures

K. Jellema reports no disclosures

## Data Availability

The data that support the findings of this study are available from the corresponding author upon reasonable request.

## Acknowlegements

M. Couman, research nurse for including patients and collecting data

## REFERENCES

1. Huisartsencijfers Volksgezondheid en Zorg [Internet]. [cited 2026 Jan 16]. Available from: https://www.vzinfo.nl/beroerte/leeftijd-en-geslacht/naar-type.

2. Vaartjes I, Reitsma JB, de Bruin A, Berger-van Sijl M, Bos MJ, Breteler MM, et al. Nationwide incidence of first stroke and TIA in the Netherlands. Eur J Neurol. 2008 Dec;15(12):1315–23. doi: 10.1111/j.1468-1331.2008.02309.x.

3. Ovbiagele B, Goldstein LB, Higashida RT, Howard VJ, Johnston SC, Khavjou OA, et al. Forecasting the future of stroke in the United States: a policy statement from the American Heart Association and American Stroke Association. Stroke. 2013 Aug;44(8):2361–75. doi: 10.1161/STR.0b013e31829734f2. Epub 2013 May 22. Erratum in: Stroke. 2015 Jul;46(7):e179. doi: 10.1161/STR.0000000000000071.

4. Arwert HJ, Groeneveld IF, Vliet Vlieland TPM, Meesters JJL. Health Care Use and Its Associated Factors 5-8 Years after Stroke. J Stroke Cerebrovasc Dis. 2019 Nov;28(11):104333. doi: 10.1016/j.jstrokecerebrovasdis.2019.104333.

5. Ayerbe L, Ayis SA, Crichton S, Wolfe CD, Rudd AG. Natural history, predictors and associated outcomes of anxiety up to 10 years after stroke: the South London Stroke Register. Age Ageing. 2014 Jul;43(4):542–7. doi: 10.1093/ageing/aft208.

6. Groeneveld IF, Arwert HJ, Goossens PH, Vliet Vlieland TPM. The Longer-term Unmet Needs after Stroke Questionnaire: Cross-Cultural Adaptation, Reliability, and Concurrent Validity in a Dutch Population. J Stroke Cerebrovasc Dis. 2018 Jan;27(1):267–275. doi: 10.1016/j.jstrokecerebrovasdis.2017.08.043.

7. van Eeden M, van Heugten C, van Mastrigt GA, van Mierlo M, Visser-Meily JM, Evers SM. The burden of stroke in the Netherlands: estimating quality of life and costs for 1 year poststroke. BMJ Open. 2015 Nov 27;5(11):e008220. doi: 10.1136/bmjopen-2015-008220.

8. Hacke W, Donnan G, Fieschi C, Kaste M, von Kummer R, Broderick JP, et al. Association of outcome with early stroke treatment: pooled analysis of ATLANTIS, ECASS, and NINDS rt-PA stroke trials. Lancet. 2004 Mar 6;363(9411):768–74. doi: 10.1016/S0140-6736(04)15692-4.

9. Almekhlafi MA, Goyal M, Dippel DWJ, Majoie CBLM, Campbell BCV, Muir KW, et al. Healthy Life-Year Costs of Treatment Speed From Arrival to Endovascular Thrombectomy in Patients With Ischemic Stroke: A Meta-analysis of Individual Patient Data From 7 Randomized Clinical Trials. JAMA Neurol. 2021 Jun 1;78(6):709–717. doi: 10.1001/jamaneurol.2021.1055.

10. Biswas R, Wijeratne T, Zelenak K, Huasen BB, Iacobucci M, Killingsworth MC, et al. Disparities in Access to Reperfusion Therapy for Acute Ischemic Stroke (DARTS): A Comprehensive Meta-Analysis of Ethnicity, Socioeconomic Status, and Geographical Factors. CNS Drugs. 2025 Apr;39(4):417–442. doi: 10.1007/s40263-025-01161-z.

11. Ikeme S, Kottenmeier E, Uzochukwu G, Brinjikji W. Evidence-Based Disparities in Stroke Care Metrics and Outcomes in the United States: A Systematic Review. Stroke. 2022 Mar;53(3):670–679. doi: 10.1161/STROKEAHA.121.036263.

12. Denny MC, Rosendale N, Gonzales NR, Leslie-Mazwi TM, Middleton S. Addressing Disparities in Acute Stroke Management and Prognosis. J Am Heart Assoc. 2024 Apr 2;13(7):e031313. doi: 10.1161/JAHA.123.031313.

13. Ford ME, Kelly PA. Conceptualizing and categorizing race and ethnicity in health services research. Health Serv Res. 2005 Oct;40(5 Pt 2):1658–75. doi: 10.1111/j.1475-6773.2005.00449.x.

14. Kim JA, Herman A, Shrader P, Alhanti B, Xian Y, Falcone GJ, et al. National Versus State-Level Racial Disparities in Acute Stroke Interventions Using Get With The Guidelines-Stroke Data. Stroke. 2025 Oct;56(10):2945–2956. doi: 10.1161/STROKEAHA.124.050446.

15. Kiefer L, Daniel D, Polineni S, Dhamoon M. Racial disparities in access to, and outcomes of, acute ischaemic stroke treatments in the USA. Stroke Vasc Neurol. 2025 Feb 25;10(1):65–70. doi: 10.1136/svn-2023-003051.

16. Emmett ES, O’Connell MDL, Pei R, Douiri A, Wyatt D, Bhalla A, Wolfe CDA, Marshall IJ. Trends in Ethnic Disparities in Stroke Care and Long-Term Outcomes. JAMA Netw Open. 2025 Jan 2;8(1):e2453252. doi: 10.1001/jamanetworkopen.2024.53252.

17. Coutinho JM, Klaver EC, Roos YB, Stam J, Nederkoorn PJ. Ethnicity and thrombolysis in ischemic stroke: a hospital based study in Amsterdam. BMC Neurol. 2011 Jun 30;11:81. doi: 10.1186/1471-2377-11-81.

18. Berkhemer OA, Fransen PS, Beumer D, van den Berg LA, Lingsma HF, Yoo AJ, et al. A randomized trial of intraarterial treatment for acute ischemic stroke. N Engl J Med. 2015 Jan 1;372(1):11–20. doi: 10.1056/NEJMoa1411587.

19. Fan S, Yang L, Ji X. Reperfusion therapy for acute ischemic stroke: Where we are and where to go. J Transl Int Med. 2025 Mar 19;13(1):1–3. doi: 10.1515/jtim-2025-0001.

20. Lee YX, Jellema K, Vlieland TPMV, van den Wijngaard IR, Hofs DECM, Arwert HJ. Ethnic disparities in health-related quality of life and cognitive function after stroke in the Netherlands. Health Qual Life Outcomes. 2026 Feb 25. doi: 10.1186/s12955-026-02498-9.

21. World Medical Association. World Medical Association Declaration of Helsinki: ethical principles for medical research involving human subjects. JAMA. 2013 Nov 27;310(20):2191–4. doi: 10.1001/jama.2013.281053.

22. von Elm E, Altman DG, Egger M, Pocock SJ, Gøtzsche PC, Vandenbroucke JP; STROBE Initiative. The Strengthening the Reporting of Observational Studies in Epidemiology (STROBE) statement: guidelines for reporting observational studies. J Clin Epidemiol. 2008 Apr;61(4):344–9. doi: 10.1016/j.jclinepi.2007.11.008.

23. Centraal Bureau voor de Statistiek (CBS). Persoon met een migratieachtergrond [internet. [cited 2026 Jan 16]. Available from: https://www.cbs.nl/nl-nl/onze-diensten/methoden/begrippen/persoon-met-een-migratieachtergrond-#:∼:text=Persoon%20van%20wie%20ten%20minste,geboren%20(de%20tweede%20generatie).

24. Feng Z, Yang M, Jin A, Ma N, Gao F, Mo D, et al. Endovascular Treatment in Patients with Large Vessel Occlusion Stroke of Different Mechanisms. Neurol Ther. 2025 Aug;14(4):1269–1283. doi: 10.1007/s40120-025-00727-9.

25. Talbot D, Massamba VK. A descriptive review of variable selection methods in four epidemiologic journals: there is still room for improvement. Eur J Epidemiol. 2019 Aug;34(8):725–730. doi: 10.1007/s10654-019-00529-y.

26. ULCLA Institute for Digital Research and Education. Exact logistic Regression. Stata Data Analysis Examples [internet]. [cited 2026 Feb 18]. Available from: https://stats.oarc.ucla.edu/stata/dae/exact-logistic-regression/.

27. Kim HY. Statistical notes for clinical researchers: Chi-squared test and Fisher’s exact test. Restor Dent Endod. 2017 May;42(2):152–155. doi: 10.5395/rde.2017.42.2.152.

28. Neuhäuser M, Ruxton GD. The Choice Between Pearson’s χ2 Test and Fisher’s Exact Test for 2 × 2 Tables. Pharm Stat. 2025 May-Jun;24(3):e70012. doi: 10.1002/pst.70012.

29. Grant SW, Hickey GL, Head SJ. Statistical primer: multivariable regression considerations and pitfalls. Eur J Cardiothorac Surg. 2019 Feb 1;55(2):179–185. doi: 10.1093/ejcts/ezy403.

30. Austin PC, Steyerberg EW. The number of subjects per variable required in linear regression analyses. J Clin Epidemiol. 2015 Jun;68(6):627–36. doi: 10.1016/j.jclinepi.2014.12.014.

31. Jackson SL, Legvold B, Vahratian A, Blackwell DL, Fang J, Gillespie C, et al. Sociodemographic and Geographic Variation in Awareness of Stroke Signs and Symptoms Among Adults – United States, 2017. MMWR Morb Mortal Wkly Rep. 2020 Nov 6;69(44):1617–1621. doi: 10.15585/mmwr.mm6944a1.

32. Govindarajan P, Friedman BT, Delgadillo JQ, Ghilarducci D, Cook LJ, Grimes B, et al. Race and sex disparities in prehospital recognition of acute stroke. Acad Emerg Med. 2015 Mar;22(3):264–72. doi: 10.1111/acem.12595.

33. Zachrison KS, Nielsen VM, de la Ossa NP, Madsen TE, Cash RE, Crowe RP, Odom EC, Jauch EC, Adeoye OM, Richards CT. Prehospital Stroke Care Part 1: Emergency Medical Services and the Stroke Systems of Care. Stroke. 2023 Apr;54(4):1138–1147. doi: 10.1161/STROKEAHA.122.039586.

34. Pirson FAVA, Boodt N, Brouwer J, Bruggeman AAE, Hinsenveld WH, Staals J, et al. Etiology of Large Vessel Occlusion Posterior Circulation Stroke: Results of the MR CLEAN Registry. Stroke. 2022 Aug;53(8):2468–2477. doi: 10.1161/STROKEAHA.121.038054.

35. Kelley RE, Buchhanolla P, Pandey A, Thapa M, Hossain MI, Bhuiyan MAN. Diagnostic yield and therapeutic implications of vascular imaging in acute ischemic stroke: prospective and consecutive study of small vessel versus large vessel ischemia. J Stroke Cerebrovasc Dis. 2025 Feb;34(2):108182. doi: 10.1016/j.jstrokecerebrovasdis.2024.108182.

36. Wolma J, Nederkoorn PJ, Goossens A, Vergouwen MD, van Schaik IN, Vermeulen M. Ethnicity a risk factor? The relation between ethnicity and large– and small-vessel disease in White people, Black people, and Asians within a hospital-based population. Eur J Neurol. 2009 Apr;16(4):522–7. doi: 10.1111/j.1468-1331.2009.02530.x.

37. Markus HS, Khan U, Birns J, Evans A, Kalra L, Rudd AG, et al. Differences in stroke subtypes between black and white patients with stroke: the South London Ethnicity and Stroke Study. Circulation. 2007 Nov 6;116(19):2157–64. doi: 10.1161/CIRCULATIONAHA.107.699785.

38. Ponce NA, Hays RD, Cunningham WE. Linguistic disparities in health care access and health status among older adults. J Gen Intern Med. 2006 Jul;21(7):786–91. doi: 10.1111/j.1525-1497.2006.00491.x.

39. Meischke H, Chavez D, Bradley S, Rea T, Eisenberg M. Emergency communications with limited-English-proficiency populations. Prehosp Emerg Care. 2010 Apr-Jun;14(2):265–71. doi: 10.3109/10903120903524948.

40. Centraal Bureau voor de Statistiek (CBS). Good command of Dutch enhances labour participation [Internet]. 08/11/2023. [cited 2026 Feb 25]. Available from: https://www.cbs.nl/en-gb/news/2023/44/good-command-of-dutch-enhances-labour-participation?

41. Rabinstein AA. Update on Treatment of Acute Ischemic Stroke. Continuum (Minneap Minn). 2020 Apr;26(2):268–286. doi: 10.1212/CON.0000000000000840.

42. Medford-Davis LN, Fonarow GC, Bhatt DL, Xu H, Smith EE, Suter R, Peterson ED, Xian Y, Matsouaka RA, Schwamm LH. Impact of Insurance Status on Outcomes and Use of Rehabilitation Services in Acute Ischemic Stroke: Findings From Get With The Guidelines-Stroke. J Am Heart Assoc. 2016 Nov 14;5(11):e004282. doi: 10.1161/JAHA.116.004282.

43. Government of the Netherlands. Standard health insurance [internet]. [cited 2026 Feb 25]. Available from: https://www.government.nl/topics/health-insurance/standard-health-insurance?

44. Ambulance-inzetten: responstijdpercentage A1-inzetten [Internet]. 2024 [cited 2026 Feb 25]. Available from: https://www.staatvenz.nl/kerncijfers/ambulance-inzetten-responstijdpercentage-a1-inzetten?

45. Pantoja-Ruiz C, Akinyemi R, Lucumi-Cuesta DI, Youkee D, Emmett E, Soley-Bori M, Kalansooriya W, Wolfe C, Marshall IJ. Socioeconomic Status and Stroke: A Review of the Latest Evidence on Inequalities and Their Drivers. Stroke. 2025 Mar;56(3):794–805. doi: 10.1161/STROKEAHA.124.049474.

